# How the Malian press treated hydroxychloroquine at the beginning of the COVID-19 pandemic

**DOI:** 10.1101/2022.07.19.22277801

**Authors:** Fabrice FE Escot, Kate KZ Zinszer, Krystelle KA Abalovi, Nathan NP Peiffer-Smadja, Abdourahmane AC Coulibaly, Adrien AS Saucier, Valéry VR Ridde

## Abstract

**Background:** The global debate on the efficacy of hydroxychloroquine (HCQ) on COVID-19 has gone far beyond the scientific framework and has been highly politicized. These issues immediately invested the debate on HCQ and made it an object of particular crystallization. This study analyzes, through the Malian press, the echo of this debate in the national background.

**Methods:** Mixed methods design, based on a review of 452 articles about COVID-19 published by six major Malian newspapers, from January 1st to July 31st 2020. Results of a content analysis with WORDSTAT8 software were further explained by a thematic qualitative analysis using and deductive-indictive approach.

**Results:** The debate on HCQ has had very little echo in the Malian press despite some interest, because of a lack of anchoring and thus of a “response” at the national level. The national health authorities, who adopted the treatment as part of clinical trials, and the press, stayed away from both the medical and the “ideological” components of the debate, despite these a priori directly involved a country like Mali.

**Conclusions:** The paper sheds light on the issues at stake in the HCQ debate based on a case study of an atypical country in terms of impacts of Covid-19. The governance of COVID helped crystallize political opposition to the presidential regime leading to a coup in August.

## INTRODUCTION

Since the beginning of diagnosing and managing the first patients with COVID-19, the question of drug treatment has been central for doctors and researchers. Faced with a severe viral lung infection, the first clinical studies focused on repositioned drugs [1]; using pre-existing drugs that could have an antiviral effect on coronaviruses in humans. The first molecules that were evaluated in this capacity were fusion inhibitors (e.g. umifenovir), RNA polymerase inhibitors (e.g. remdesivir, ribavirin), protease inhibitors (e.g. lopinavir/ritonavir) as well as endosomal acidification inhibitors (e.g. chloroquine, hydroxychloroquine) [2].

Among these molecules, hydroxychloroquine was by far the most evaluated drug in the initial phase of the pandemic with, at the end of April 2020, more inclusions of patients in trials evaluating hydroxychloroquine than for all other antiviral drugs combined.[3] (Hydroxy)chloroquine is a very accessible antimalarial drug that has been widely used by clinicians to treat COVID-19, despite the lack of evidence to support its use. The very rapid rise in interest in (hydroxy)chloroquine can be easily connected to its promotion at the end of February 2020 by Professor Raoult in France, which was subsequently supported by many populist politicians including, among others, Donald Trump and Jair Bolsonaro in March 2020[1]. Professor Raoult claims were first based on the sharing of preliminary clinical data by Chinese teams in February 2020, the data of which were never published, and then on biased non-comparative clinical studies [4][5][6]. Azithromycin (an antibiotic) was then added without clinical evidence to the treatment “protocol” recommended by Raoult’s team in France. Although these two molecules were found to have an in vitro effect on SARS-CoV-2 on animal cell lines, like many molecules in the pharmacopoeia, it was quickly found that this effect was not reproduced in human cells [7]. These findings were quickly confirmed in animal models, for example in macaques infected with SARS-COV-2 [8] which develops a COVID-19 pneumonia close the one found in humans.

However, despite these results leaving little hope of clinical efficacy, the evaluation of the molecule in humans has continued in numerous randomized therapeutic trials evaluating the efficacy and safety of hydroxychloroquine in COVID-19. None of these large-scale trials (Recovery, DisCoVeRy, Solidarity trials, etc.) found any efficacy of hydroxychloroquine in COVID-19, regardless of the dose used, the association with other molecules including azithromycin, or the phase of the disease.

The lack of efficacy of HCQ for COVID-19 has been confirmed in large meta-analyses gathering data from tens of studies [9] [10] and is now consensus in the medical and scientific community. None of the scientific agencies or medical societies with recent and up-to-date therapeutic recommendations currently advise the use of hydroxychloroquine in the prevention of SARS-CoV-2 infection or in patients with COVID-19.

Mali is a West African Sahelian country of twenty million inhabitants in 2021, affected late (the first two cases were reported on March 24, 2020) by the pandemic (Figure 1). Incidence rates have been lower than those reported by many other countries (approx. 3%), even African countries, notably Senegal [11]. Senegal, a border country of seventeen million inhabitants in 2021, has suffered a higher number of declared cases than other West African countries.

**Figure 1:**
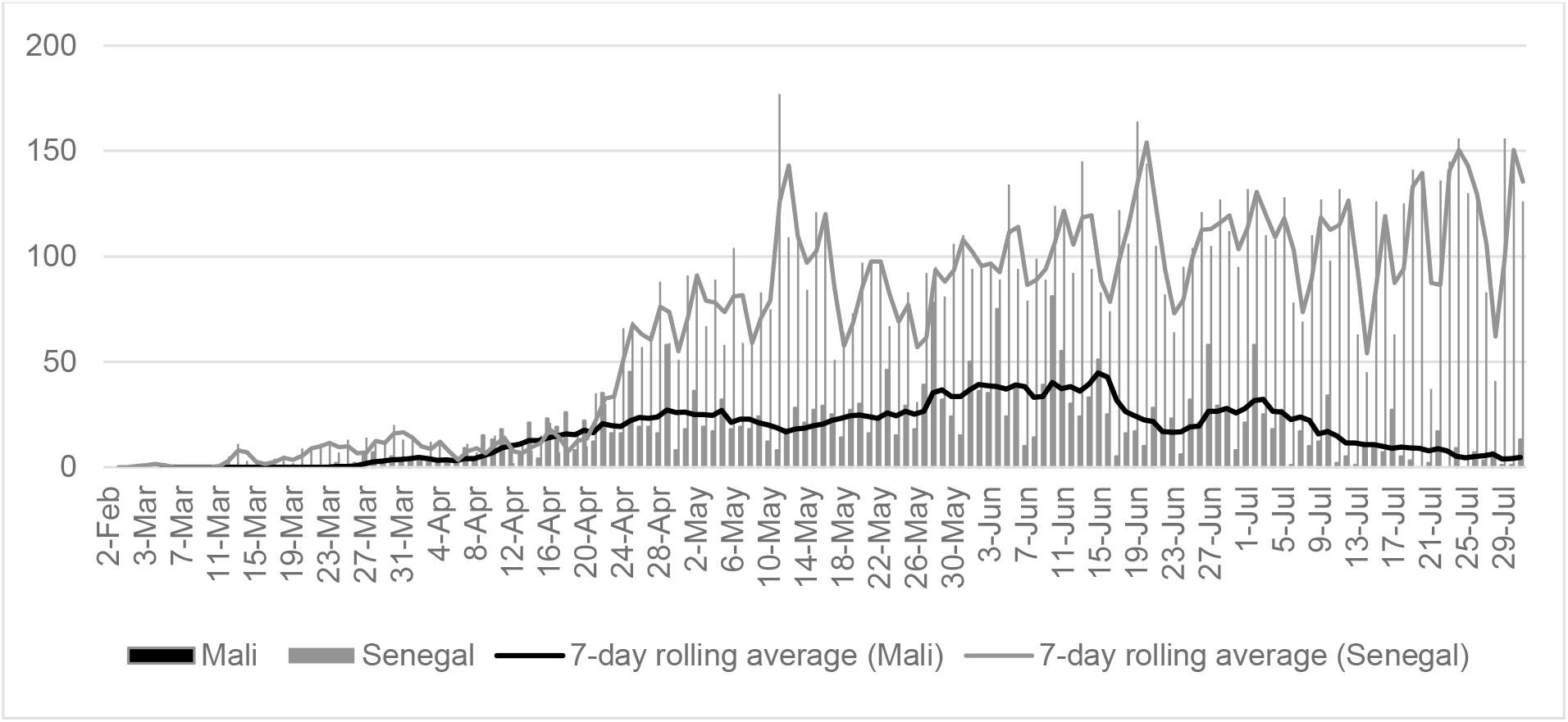
Daily total of confirmed SARS-CoV-2 cases in Mali and Senegal and 7-day rolling average, from 28 February to 31 July 2020 Source: https://www.covid19afrique.com Legend: The total length of the bars represents the incidence over the months. The line graphs represent the 7-day rolling average. The upper line and associated bars, thinner and in grey, represent the data for Senegal, the lower one with larger bars and in black, the data for Mali.

Clinical trials on HCQ conducted by Prof. Seydi, a Senegalese infectiologist, led him to defend its efficacy, including an inconclusive article published in June 2021 [12].

In Mali, the COVID-19 treatment protocol was adopted on April 1, 2020 by the Ministry of Health, four days after the first reported cases. The treatment includes chloroquine phosphate tablets combined with Azythromicin and vitamin C, plus for asymptomatic patients Xarelto and for symptomatic patients Levonox, Solumedrol, and Betacef or Levofloxacin.

In this controversial and pandemic context, the objective of this article was to measure and understand how the press in Mali has addressed the issue of treatment against COVID-19 and in particular the use of hydroxychloroquine. The results of this study will be useful, despite that the Malian press is not widely read by the general population outside of Mali, it remains an important source of information for policy makers in Mali and in West Africa, and therefore has the power to influence political and clinical decision making [13].

## METHOD

### Design

This study was based on a sequential explanatory design [14], meaning that the quantitative results informed the qualitative methods. The qualitative data were analyzed to interpret some of the quantitative results and to provide in-depth understanding on how the Malian press addressed HCQ specifically.

Until 2020, the Malian press, like other West African countries, consisted of a large number of private newspapers, with the exception of the state-owned daily newspaper, which benefited from a certain degree of freedom of the press. Many newspapers (daily and mostly periodical) focused on national politics. The online press has developed strongly over the past 20 years, particularly through the main generalist newspapers.

### Population

An empirical selection of newspapers was based on previous media studies [15]. All articles on COVID-19 were systematically collected from three exclusively online press sites (Maliweb.net, Maliactu.net, and ORTM.com) and from three newspaper websites that also publish in print: L’Essor (State-owned daily newspaper), L’Indépendant (daily newspaper), and Le Journal du Mali (JDM - weekly newspaper).

### Sample

Articles were searched from 01/01/2020 to 07/31/2020 and were manually identified with keywords: COVID, COVID-19 and Coronavirus, in the title or the full text. The full text from each identified article was manually extracted into a Word file, including identifiers such as source, date, title and authors, and the identifier were also managed in an Excel file. We estimate that the medias under study publish around 600 per week.

### Descriptive analysis

The text of the articles was uploaded into WORDSTAT8 for descriptive and content analyses how COVID-19 treatments (including hydroxychloroquine) were approached. With the *categorization* function in WORDSTAT8, we identified words related to treatment of COVID-19 in the articles using manual review and the WORDSTAT8’s “suggest” function for synonyms and similar words. We classified the words into the following categories: generic terms, mechanical treatments, chloroquine and hydroxy-chloroquine, other synthetic drugs, and plant-based treatments (Table 1). We then used the *dendrogram* function (concept clustering), to investigate terms associated with words chloroquine and hydroxychloroquine in the articles.

**Table 1:** Words classified into treatment categories and occurrences.

| Categories |  | Number of occurrences<br>(88<br>“treatment<br>” articles) |  | Words | Number of occurrences |  |
| --- | --- | --- | --- | --- | --- | --- |
|  |  |  |  |  | 88<br>“treatment<br>” articles | 17<br>“HCQ”<br>articles |
| Generic words |  | 181 |  | Treatment | 93 | 39 |
|  |  |  |  | Drug(s) | 88 | 45 |
| Specific words | (Hydroxy-) chloroquine | 141 | 60 | Chloroquine | 36 | 36 |
|  |  |  |  | Hydroxychloroquine | 24 | 24 |
|  | Mechanical treatments |  | 59 | Respirator | 21 | 6 |
|  |  |  |  | Resuscitation | 20 | 4 |
|  |  |  |  | Oxygen | 12 | 4 |
|  |  |  |  | Oxygen extractor(s) | 2 | 0 |
|  |  |  |  | Oxygen concentrators | 1 | 0 |
|  |  |  |  | Multi-parameter monitors | 1 | 0 |
|  |  |  |  | Source(s) of oxygenation | 1 | 0 |
|  |  |  |  | Mechanical ventilation | 1 | 0 |
|  | Other drugs |  | 12 | Azythromycin | 6 | 6 |
|  |  |  |  | Corticoids | 3 | 2 |
|  |  |  |  | Antibiotic | 1 | 1 |
|  |  |  |  | Lopinavir | 1 | 1 |
|  |  |  |  | Ritonavir | 1 | 1 |
|  | Plant-based treatments |  | 10 | Plant | 5 | 4 |
|  |  |  |  | COVID Organics | 4 | 2 |
|  |  |  |  | Avipirin | 1 | 1 |

The first two columns of text indicate the categories, the first columns of numbers the number of occurrences of words in each category in the 88 articles addressing the treatment. The third column of text indicates the words used, the last columns of numbers the number of occurrences of words in each category, respectively in the 88 articles dealing with treatment and in the 17 articles dealing specifically with HCQ.

### Qualitative analysis

Articles with at least one mention of the term chloroquine or hydroxychloroquine were manually extracted from the set. These articles were manually analyzed using a hybrid deductive-inductive approach to identify the sources, the themes addressed, the vocabulary used and the arguments developed to justify, where appropriate, the position of the actors [16].

The integration of the results was done in the discussion section as recommended for a mixed methods design [14].

## FINDINGS

### Treatments and HCQ

The 88 articles that contained concepts of COVID-19 treatment constituted 19% of the articles identified on COVID-19 (n = 452). The 20 terms of interest represent 2% of the approximate 1,000 words of interest identified by WORDSTAT8 and 0.5% of the corresponding approximate total of 60,000 occurrences.

The most frequently cited treatment categories in the selected articles were generic words (181 occurrences). (Hydroxy-) chloroquine was the second category with 60 occurrences. Other categories were mechanical treatments (59), other “chemical” drugs (12) and plant-bases treatments (10). The five categories identified include 20 words, of which 7 provided 294 occurrences or 90% of the total (Table 1).

The 452 articles in the corpus have an average of 530 words, the 17 “HCQ” articles have an average of 890 words, and also concentrate a significant part of the occurrences of the other words related to the treatment of COVID-19 identified in the whole corpus (Table1).

The appearance of treatment in the media began in January 2020 (related with the situation in China), increased in frequency with the appearance of first COVID-19 cases in Mali (March 27) and especially when incidence of COVID-19 cases was the highest (up to 84 cases per day in June 2020) (Figure 1).

The appearance of hydroxychloroquine in the media began with the first COVID-19 cases in Mali (March 27). The highest frequencies indicated in March, May and July 2020 correspond to international news, including the adoption of hydroxychloroquine by France in March and the publication of clinical studies by The Lancet in May and July 2020 [17] (Figure 2).

**Figure 2:**
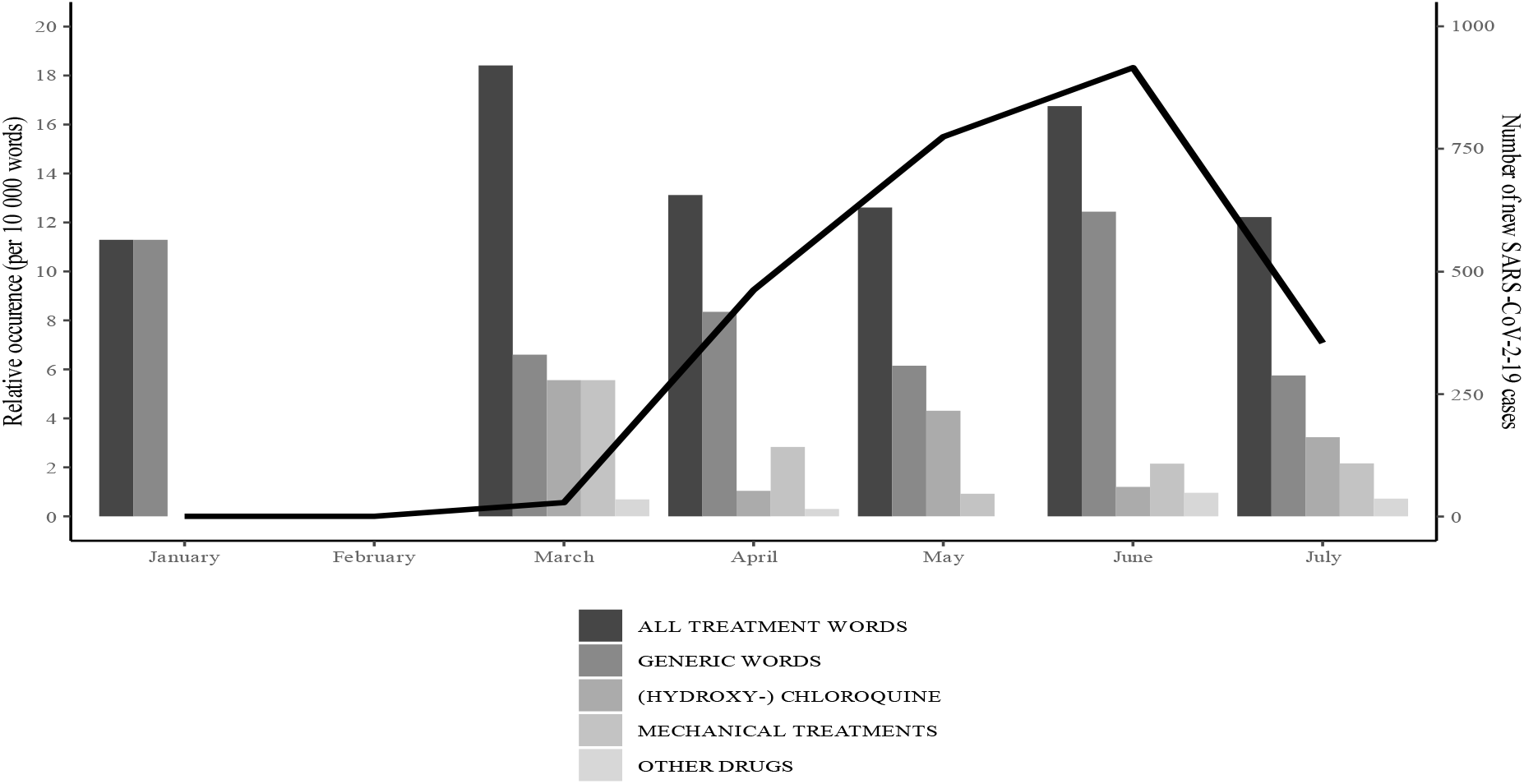
Relative occurrences of the four most frequently reported treatment categories in the Malian press by month (left axis) and monthly cases of SARS-CoV-2 in Mali (right axis) Legend: The total length of the bars represents the relative occurrence per 10,000 words of each word category over the months of January to July 2020. The line graph represents the number of new cases in Mali per month.

Analyzing co-occurrences through hierarchical clustering shows that hydroxychloroquine, unlike chloroquine, was more discussed in the context of studies (Figure 3). Clinical trials on *Covid organics*, a Malagasy artemisia-based treatment, were also mentioned (n=2).

**Figure 3:**
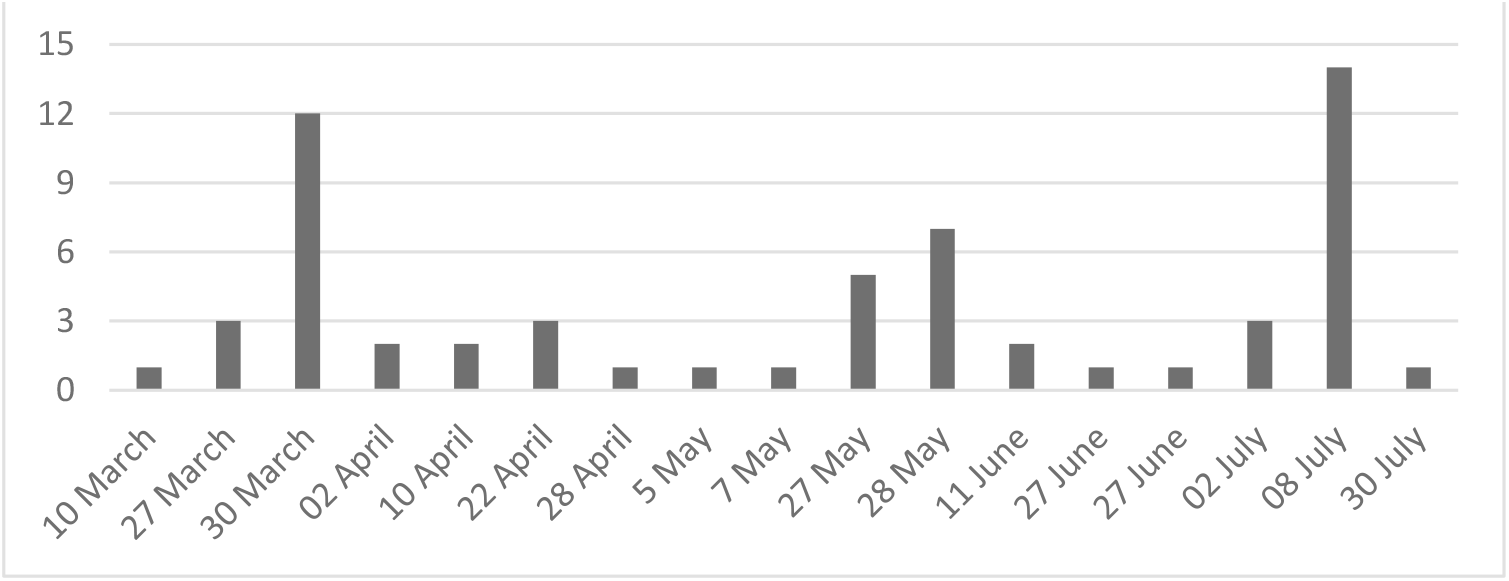
Occurrences of the (H)CQ word for each of the 17 articles (in chronological order of publication, from March to July 2020. Legend: The total length of the bars represents the number of occurrences of (hydroxy-)chloroquine per article.

**Figure 4:**
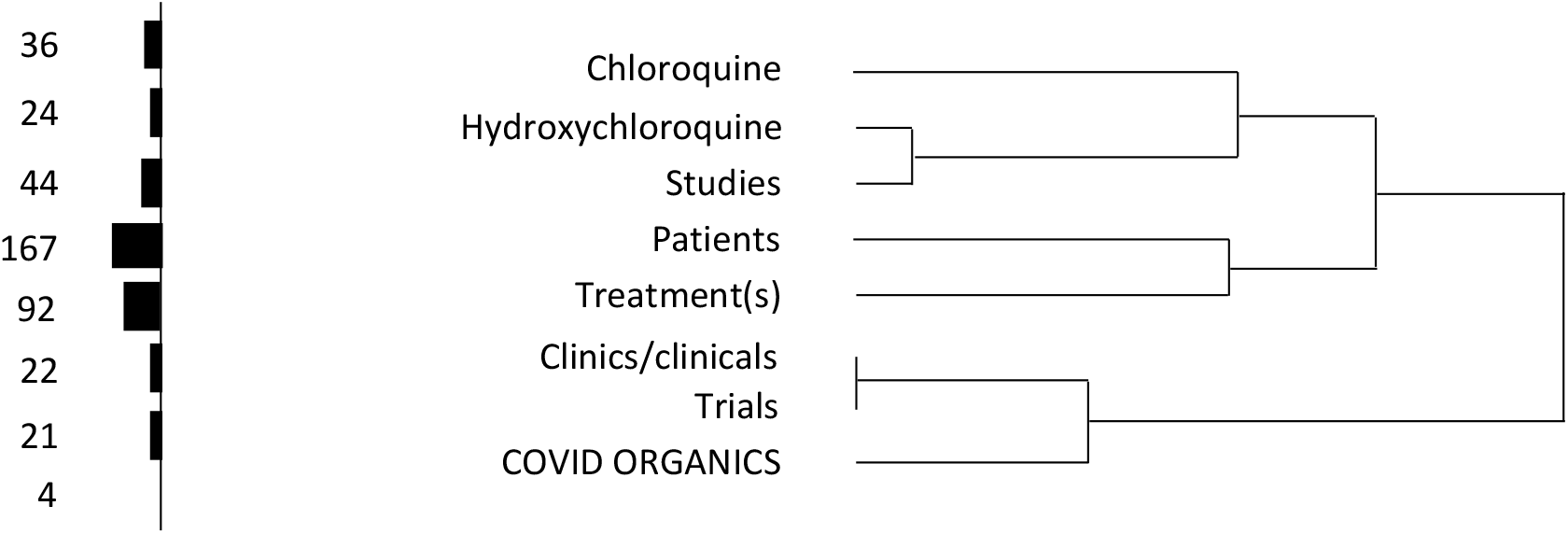
Hierarchical cluster on the “Treatment” subject automatically detected by WORDSTAT8 software and occurrences of the related words. Legend: The hierarchal clustering of terms as indicated by lines and also the frequency of occurrence of the different terms indicated by the bar chart on the left.

### The press and HCQ efficacy

None of the 17 articles discuss in-depth the treatment protocol adopted in Mali on 1 April 2020. However, four articles mention HCQ with one briefly describing the treatment.

> “*Chloroquine phosphate is given for 10 days and dosed at 500mg every 12 hours. Azithromycin 500mg on day 1 and 250mg on days 2-5. And vitamin C is given twice for 5 days*. [L’Essor, June 11, 2020]

The international debate on the clinical effectiveness of HCQ was the subject of 8 articles (46 occurrences), most of them relaying the views of national and international health experts. These articles concentrate on the use of the term “hydroxychloroquine” and references to treatments associated with HCQ (lopinavir, ritronavir, azythromycin, antibiotics) or alternative treatments (avipirine, COVID-Organics/artemisia).

The press reported the hopes raised by Raoult’s statements, relayed by African doctors in the media. These experts cited, as of March-April 2020, the low number of deaths among the cases treated in African health structures as conclusive results of the effectiveness of HCQ. They also denounce a dramatization of the coronavirus in the Sahelian context. However, the press also reports on the lack of scientific consensus and the clinical trials underway. The articles reiterate the need for treatment under medical supervision, the contraindications, and the risks associated with self-medication.

> *“With the hope raised by chloroquine in the treatment of the conoravirus in recent times, our compatriots are jostling each other at the gates of the pharmacies to obtain this molecule. Some see it as a panacea and do not hesitate to self-medicate. [Far be it from us to engage in a sterile debate on the effectiveness or otherwise of chloroquine. This debate is conducted at other levels with scientific arguments for and against. Moreover, the World Health Organisation (WHO) has not approved the thesis on the effectiveness of chloroquine in the treatment of Covid-19.”*, L’Essor, March 30, 2020]

Following the suspension of the clinical trials by the WHO, the Malian Minister of Health, without taking a formal position on the therapeutic protocol, put the health impact of the epidemic in Africa into perspective and declared himself open to other treatments. On the other hand, the Malian press mentions Professor Seydi’s team (who conducted trials on HCQ in Senegal[12]), which affirms on this occasion, and again in July, that Senegal will maintain its therapeutic protocol, considering that clinical studies have provided positive results. On 8 July 2020, following the contradictory study published by The Lancet, Maliweb devoted a retrospective article to the debate. It closes with the uncertainties:

> *“A new twist in the debate on the effectiveness of hydroxychloroquine against Covid? A study conducted in China and published on 3 July in the famous scientific journal The Lancet, would prove that the level of Covid 19 would be lower in patients taking hydroxychloroquine. The Lancet’s “backtracking”, as stated by the europe-israel.org website, has been shared by thousands of people on social networks. [*…*] Although clinical trials have resumed, the effectiveness of the Covid treatment is still being debated. [“In short, the miracle cure for Covid has not yet arrived.”* Maliweb, July 08, 2020]

### Anxiety about the problems of supplying health facilities with HCQ

As early as 2 April 2020, concerns were made over having adequate supply of hydroxycholoroquine (L’Indépendant, April 2). In early May, the weeklies Le Témoin and Le Journal du Mali denounced the shortages affecting health staff and patients in “an inhospitable and criminal service” (Le Témoin, May 5). L’Essor recalls that the problems stem from the global shortage of medicines and equipment, and from the pressure of the international pharmaceutical market, which was negatively impacted by border closures, and financially weak African countries were further disadvantaged. Several articles published by the Journal du Mali, Maliweb and Maliactu praise Morocco, particularly in June-July 2020, for having supplied fifteen African countries with, among other things, 75,000 boxes of chloroquine and 15,000 boxes of Azythromycin. However, they warn of future challenges and competition between African countries in accessing medical inputs (drugs and medical supply).

> *« The AU Commission is calling on member states to strengthen their capacity to manufacture medical products. [*…*] On a global level, the harshness of the health crisis has somewhat undermined solidarity with nations that are fighting alone without concern for the fate of their neighbours.” [“Covid-19* : *le Maroc en tête de la riposte africaine”*, Le Journal du Mali, June 27, 2020]

## DISCUSSION

Despite the adoption of the controversial treatment of COVID-19 with HCQ by the Malian government, the arguments against its use were not the subject of debate in the press nor in official/institutional communications. Interestingly, other aspects of the national response plan were communicated through press releases or in the media: collective prevention measures (border closures, curfew, physical distancing) and, for symptomatic cases, on the setting up of care structures and their equipment with mechanical treatment [11].

In the face of international scientific controversy [18–22], the Malian state was faced with a choice that was fraught with ideological drift [18–20], in an African and national context that was clearly in favor of hydroxychloroquine, making it difficult to communicate an unambiguous message [21–23]. The domestic political context, with a very critical opinion of the State itself and in particular of its governance of the pandemic [11], certainly had an influence in making it impossible to make the political choice of refusing a “solution of hope for the Malian population”[20,23–25]. During 2020, there was growing opposition in Mali to President Keita, not least because of the endemic corruption. The rift between the presidency and Malian society led to the creation of a frontal opposition collective and to mass demonstrations (June and July 2020). Finally, on 18 August 2020, a coup d’état brought a military junta to power. The country has therefore taken (and defended) a position that is both hopeful and empowering for Africa [24] and has integrated HCQ into its medical protocol [23–25], in the spirit of the “duty to care” invoked by Professor Raoult [19]. At the same time, the Minister of Health (former UNAIDS Deputy Executive Director, close to France) never rejected the possibility to adopt the African plant-made alternative drugs, but in both matters declared to comply to the WHO norms. This may suggest that the Malian state never thought it had a real treatment, unlike Senegal, which claimed conclusive results and published an article of questionable quality [12].

Although the Malian press had been aware of the debate of HCQ effectiveness, particularly in France and then worldwide, it has not taken a position. Indeed, given the absence of a clear national directive, it did not have the capacity to take a formal position, feeling that it was out of their area of expertise. This led to relaying the available information, beginning with Pr Raoult at the beginning, African doctors, WHO decisions, and followed by the Lancet publications. The Malian press often used articles from the foreign media, particularly French media (RFI, AFP, France 24). This cautious approach and the few articles published on the subject cannot be attributed to the economic impact of the pandemic on the press. Indeed, many newspapers continued to publish regularly throughout the period. Nor can this circumspective attitude be the result of manipulation of the press by the political authorities. On the contrary, the press has not ceased to criticize the governance of the pandemic and the measures taken by the state. This distinguishes the highly critical West African press [24,26] from the East African press, which has experienced decreased freedom during the pandemic [27].

Nevertheless, the caution of health institutions about the virtues of chloroquine and that of the press did not prevent abuses. The population was exposed to the messages about chloroquine, with self-medication practices facilitated by informal and sometimes illegal market mechanisms resulting in the circulation of non-compliant (simple chloroquine) or counterfeit medicines [21,23,25,28]. In this context, the press discourse reflected a social acceptance of HCQ as a preferred solution. This “default” approach by the health authorities and the press, combined with the dissemination of fake news [23], reinforced public expectations and anxieties about HCQ supply.

The end of the study period (July 2020) is a methodological limitation as the HCQ debate was not yet finished. In August 2020, following the publication of the results of the Solidarity trials, the WHO definitively withdrew chloroquine from the protocols (ref). However, some countries and amongst them African countries have kept hydroxychloroquine, some in a very assertive way (Algeria, Brazil, Senegal). This is also the case in Mali, but still without any official discourse and without an assertive stance. Six press articles about HCQ were published by these newspapers from August 2020 to March 2021 (refs), notably about these results and the debate “WHO/Raoult-Seydi” – one of them indicating that HCQ was still used by the health system. An empirical study on the clinical realities of the use of chloroquine therefore remains to be conducted in the health facilities in order to better understand the lasting practice of HCQ [9].

## Data Availability

All data produced in the present study are available upon reasonable request to the authors

## References

1. Sattui SE, Liew JW, Graef ER et al. Swinging the pendulum: lessons learned from public discourse concerning hydroxychloroquine and COVID-19. Expert Review of Clinical Immunology 2020;16:659–66.

2. Fragkou PC, Belhadi D, Peiffer-Smadja N et al. Review of trials currently testing treatment and prevention of COVID-19. Clinical Microbiology and Infection 2020;26:988–98.

3. Peiffer-Smadja N, Lescure F-X, Sallard E et al. Anticovid, a comprehensive open-access real-time platform of registered clinical studies for COVID-19. Journal of Antimicrobial Chemotherapy 2020;75:2708–10.

4. The RECOVERY Collaborative Group. Effect of Hydroxychloroquine in Hospitalized Patients with Covid-19. N Engl J Med 2020;383:2030–40.

5. Ader F, Peiffer-Smadja N, Poissy J et al. An open-label randomized controlled trial of the effect of lopinavir/ritonavir, lopinavir/ritonavir plus IFN-β-1a and hydroxychloroquine in hospitalized patients with COVID-19. Clinical Microbiology and Infection 2021;27:1826–37.

6. Mitjà O, Corbacho-Monné M, Ubals M et al. Hydroxychloroquine for Early Treatment of Adults With Mild Coronavirus Disease 2019: A Randomized, Controlled Trial. Clinical Infectious Diseases 2021;73:e4073–81.

7. Hoffmann M, Mösbauer K, Hofmann-Winkler H et al. Chloroquine does not inhibit infection of human lung cells with SARS-CoV-2. Nature 2020;585:588–90.

8. Maisonnasse P, Guedj J, Contreras V et al. Hydroxychloroquine use against SARS-CoV-2 infection in non-human primates. Nature 2020;585:584–7.

9. Fiolet T, Guihur A, Rebeaud ME et al. Effect of hydroxychloroquine with or without azithromycin on the mortality of coronavirus disease 2019 (COVID-19) patients: a systematic review and meta-analysis. Clinical Microbiology and Infection 2021;27:19–27.

10. Singh B, Ryan H, Kredo T et al. Chloroquine or hydroxychloroquine for prevention and treatment of COVID-19. Cochrane Infectious Diseases Group (ed.). Cochrane Database of Systematic Reviews 2021;2021, DOI: 10.1002/14651858.CD013587.pub2.

11. Bonnet E, Bodson O, Le Marcis F et al. The COVID-19 pandemic in francophone West Africa: from the first cases to responses in seven countries. BMC Public Health 2021, DOI: https://doi.org/10.1186/s12889-021-11529-7.

12. Taieb F, Diallo Mbaye K, Tall B et al. Hydroxychloroquine and Azithromycin Treatment of Hospitalized Patients Infected with SARS-CoV-2 in Senegal from March to October 2020. 2021.

13. Escot F. La presse et la protection sociale au Mali. In: Ridde V (ed.). Vers Une Couverture Sanitaire Universelle En 2030 ?. Québec: Science et bien commun, 2021.

14. Pluye P, Hong QN. Combining the Power of Stories and the Power of Numbers: Mixed Methods Research and Mixed Studies Reviews. Annu Rev Public Health 2014;35:29–45.

15. Escot, F., Ousseini A. Presse sous influence, presse sans opinion. Traitement des politiques de gratuité des soins par les presses écrites nigérienne et malienne. In: Olivier de Sardan J-P, Ridde V (eds.). Une Politique Publique de Santé En Ses Contradictions. La Gratuité Des Soins Au Burkina Faso, Au Mali et Au Niger. Paris: Khartala, 2014.

16. Huberman AM, Miles MB. The Qualitative Researcher’s Companion. Thousand Oaks, CA: Sage Publications, 2002.

17. Mehra MR, Desai SS, Ruschitzka F et al. RETRACTED: Hydroxychloroquine or chloroquine with or without a macrolide for treatment of COVID-19: a multinational registry analysis. The Lancet 2020:S0140673620311806.

18. Schultz E, Ward JK. Science under Covid-19’s magnifying glass: Lessons from the forst months of the chloroquine debate in the French press. Journal of sociology 2021, DOI: DOI: 10.1177/1440783321999453.

19. Sauvayre R. Retour sur le débat médiatique et éthique concernant le traitement contre la Covid-19 à base de chloroquine. Les Cahiers de l’Espace Ethique 2020, DOI: hal-02965645.

20. Peiffer-Smadja N, Rebeaud ME, Guihur A et al. Hydroxychloroquine and COVID-19: a tale of populism and obscurantism. Lancet Infet Dis 2021, DOI: doi: 10.1016/S1473-3099(20)30866-5.

21. Cunliffe-Jones P, Diagne A, Gaye S et al. Misinformation Policy in Sub-Saharian Africa: From Laws and Regulations to Media Literacy. University of Westminster Press 2021, DOI: https://doi.org/10.16997/book53.

22. Ioannidis JP, Tezel A, Jagsi R. Overall and COVID-19-specific citation impact of highly visible COVID-19 media experts: bibliometric analysis. BMJ Open 2021;11, DOI: doi:10.1136/bmjopen-2021-052856.

23. Desclaux A. La mondialisation des infox et ses effets sur la santé en Afrique : l’exemple de la chloroquine. The Conversation 2020.

24. Jaffré Y, Hane F, Kane H. Une épreuve de dignité : regard anthropologique sur les réponses à la Covid-19 en Afrique de l’Ouest. Alterntives Humanitaires 2020:86–113.

25. White NJ, Watson JA, Hoglund RM et al. COVID-19 prevention and treatmnent: A critical analysis of chloroquine and hydroxychloroquine clinical pharmacology. Plos Med 2020, DOI: https://doi.org/10.1371/journal.pmed.1003252.

26. Bisson L, Schmauder A, Claes J. The politics of COVID-19 in the Sahel. 2020.

27. RSF. Covid makes African journalism more vulnerable than ever. 2021.

28. Ogunleye OO, Basu D, Mueller D et al. Response to the Novel Corona Virus (COVID-19) Pandemic Across Africa: Successes, Challenges, and Implications for the Future. Frontiers in Pharmacology 2020;11:1205.

